# Applying Human-Centered Design and Iterative Programming to Identify and Reach Zero-Dose and Under-Immunized Children in Kenya: Lessons from the Zero-Dose Learning Agenda

**DOI:** 10.64898/2026.09.01.26361953

**Authors:** Christopher Obong’o, Shamim Omar, Stella Wanjiru, Dickson Otiangala, Quinter Tabu, Mouline Ochieng’, Edith Jepleting’, Grace Njenga, Brian Taliesin

## Abstract

**Background:** Despite decades of investment in immunization, an estimated 14.3 million children worldwide remain zero-dose (ZD). Kenya’s national coverage of approximately 72% masks significant subnational disparities, with existing methods failing to capture the context-specific drivers of zero-dose status.

**Objective:** To apply human-centered design (HCD) and continuous learning to understand ‘Who’ are ZD children, ‘Why’ they are ZD and to co-design and pilot tailored interventions in two high-burden Kenyan sub-counties.

**Methods:** We implemented the Zero-Dose Learning Agenda in Turkana Central (rural, nomadic) and Rachuonyo North (semi-urban) between November 2023 and December 2025. We applied PATH’s Living Labs ‘4D’ HCD methodology – Discover, Define, Dream, Design – to guide the data collection process (interviews, observations, focus group discussions [FGDs], facilitated discussions), analysis (quantitative surveys, qualitative and data triangulation), co-creation/ideation (ideation, concept validation), prototyping (detailed concept sheet development, measurement and implementation plans) and iterative testing of three ZD interventions in select facilities and communities.

**Results:** We engaged 798 participants and identified six distinct caregiver archetypes per site. Gender norms dominated drivers in Rachuonyo North; geographic access and marginalization dominated in Turkana Central. Chanjo Talks Kazini (n=188) achieved 60.1% completion and 92.5% satisfaction; perceived spousal support increased from 37.1% to 74.4%. Quality Household Assessment and Sensitization trained 68 community health promoters, reached 874 households, and identified 15 zero-dose and 39 under-immunized children, all subsequently vaccinated. Chanjo Mashinani conducted 30 low-cost outreach events, reached 266 zero-dose children, and reduced costs by over 70% versus conventional outreach. Iterative adaptation uncovered barriers including male caregivers’ fear of HIV testing, invisible to baseline instruments.

**Conclusion:** Combining human-centered design with continuous learning enables immunization programs to diagnose context-specific barriers and correct course in real time. Zero-dose children share a label but not pathways to exclusion; effective reach requires tailored strategies that account for local gender dynamics, health system trust, and access constraints.

**Teaser key message:** ZD and UI children require tailored approaches and interventions to be identified, understood and reached, all while ensuring local voices engagement are not left at problem identification, and resources are availed to support weaker health systems.

## INTRODUCTION

### The Global Zero-Dose Challenge

Despite decades of investment in immunization programs, an estimated 14.3 million children worldwide remain zero-dose (ZD)—having received no routine vaccinations—and an additional significant proportion are under-immunized (UI), having started but not completed their vaccination schedules ^1^. The Immunization Agenda 2030 has prioritized reaching these missed children as essential to reducing vaccine-preventable disease burden and achieving universal health coverage ^2^. However, zero dose children (ZDC) are not a homogeneous population; they are concentrated in specific geographic, social, and economic contexts that demand tailored approaches rather than one-size-fits-all programmatic solutions ^3^.

### The Kenyan Context

Kenya presents a particularly complex ZD landscape. Kenya’s national immunization coverage for children under one year stands at approximately 72%, ^4^ yet this aggregate figure masks significant sub-national heterogeneity. ZDC are sparsely distributed across most of the country but concentrated in specific hard-to-reach locations, particularly arid and semi-arid lands and certain lake basin communities ^5^.

The Kenya National Vaccines and Immunization Program (NVIP), has prioritized reaching all vaccine-eligible children. However, existing identification methods are insufficiently granular, often failing to capture the “why” behind ZD status or the diverse profiles of missed children and their caregivers ^6^. Interventions to reach ZD children have historically been generalized and designed through top-down approaches, insufficiently tailored to local drivers and contexts ^6^, ranging from Big Catch-Up campiagns to reach those not previously immunized and local community stakeholder engagements to generate demand.

#### Box 1

**Zero-Dose Learning Agenda: a 2-year initiative to better understand how to reach and reduce zero-dose children**

The Zero-Dose Learning Agenda (ZDLA) was a 2-year learning initiative by the Gates Foundation to better understand how to accelerate progress toward the IA2030 goal of achieving a 50% reduction in ZD. Through ZDLA, partners applied learn-by-doing (LxD) techniques and practices in high-ZD settings to:

- Diagnose drivers of ZD and understand their root causes;
- Work collaboratively with end-users, including caregivers, community leaders, healthcare workers, and government officials, to co-design, implement, and iteratively adapt interventions;
- Reflect gender-related barriers and responsiveness; and,
- Capture costing information.

This two-year initiative (2023–2025) operated across select subnational areas in DRC, Ethiopia, India, Kenya, Nigeria, and Pakistan.

The overall LxD approach applied in ZDLA connects Human-Centered Design (HCD) with continuous learning. LxD is a general term for a set of approaches that can be used to better understand and address persistent health system challenges, supporting more flexible, efficient, and equitable health service delivery. HCD is a collaborative and empathetic approach to designing solutions whose tools and processes can be applied to any problem, including in the field of global health. HCD helps us think differently about problems, resulting in more effective solutions by prioritizing listening and empowering users – those ‘influencing’ or ‘impacted by’ a problem – through a process led by in-country experts ^6^.

LxD embeds HCD, adaptive management or continuous quality improvement, and gender-responsive practices in routine health system workflows to improve processes, products or tools, and planning at different levels of the health system.

This paper describes the implementation of ZDLA in Turkana Central (TC) and Rachuonyo North (RN) sub-counties between November 2023 and December 2025 with the aims of understanding:

-Who are ZD children?
-Why are they ZD?
-What context specific innovative solutions can help reach ZDC?

## METHODS

### Setting and Site

In collaboration with NVIP, we purposively selected two sub-counties representing diverse ZD contexts, TC, in northern Kenya, represents rural, nomadic, hard-to-reach communities with 60% immunization coverage—12 percentage points below national rate—and RN, in western Kenya, represents a semi-urban, more static population bordering Lake Victoria with distinct challenges related to adolescent pregnancies, gender dynamics, and health worker attitudes and 76% coverage ^5^.

**Table 1:** Profile of sites.

| Turkana Central <sup>5</sup> | Rachuonyo North <sup>5</sup> |
| --- | --- |
| • Low population density – 13 persons/km<sup>2</sup><br>• 82% poverty prevalence<br>• ~20% literacy<br>• 60% immunization coverage<br>• 51% home deliveries | • Population density – 410 persons/km<sup>2</sup><br>• ~55% literacy<br>• High adolescent pregnancy (23%)<br>• 76% immunization coverage |

### The 4D HCD Approach

To operationalize the LxD approach in these contexts, we applied an adapted version of PATH Living Labs’ user-led ‘4D HCD’ approach—Discover, Define, Dream, Design—with two additional phases (Deliver and Adapt) corresponding with continuous learning to keep interventions responsive to emerging realities.

**Figure 2.**
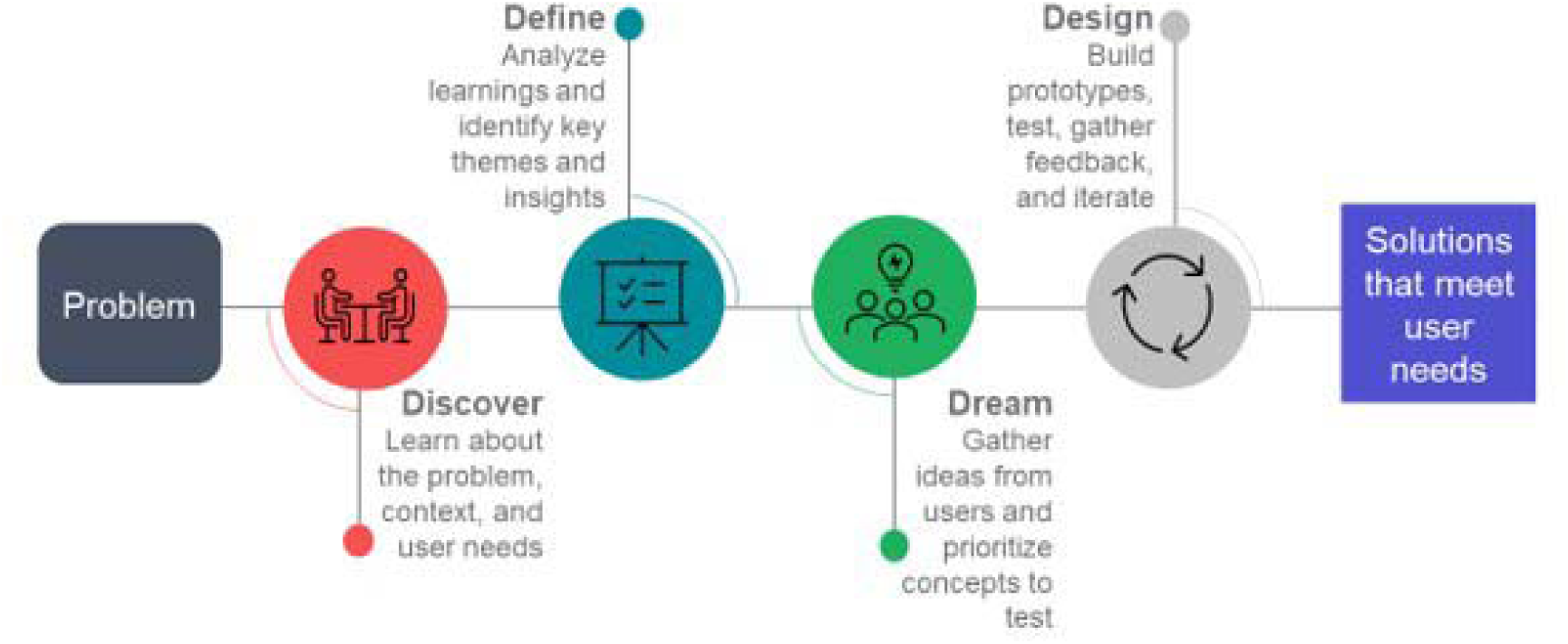
PATH’s Living Labs ‘4D’ HCD approach

In **Discover,** community health promoters (CHPs) administered household screening tools in RN and facility-based line-listing and outreach camp screening in TC to map and identify ZDC and their caregivers. To further understand their reasons for being ZDC, we conducted rapid qualitative research including observations, in-depth interviews, and focus group discussions with iterative sampling until thematic saturation was achieved. We triangulated findings with secondary data comprising national, county, and sub-county reports and a survey of ZD households. We applied the Exemplars of Global Health (EGH) framework ^7^ and adapted UNICEF’s Journey to Health & Immunization map ^8^ to guide development of data collection tools and categorization of findings.

#### Box 2

**Exemplars of Global Health (EGH) Framework**

The EGH framework conceptually describes the drivers of vaccination across the spectrum of demand and supply by categorically placing drivers under either intent, access or readiness, where ‘Intent’ looks at how awareness and knowledge, attitudes and perceptions, norms, and agency contribute to caregiver decision-making and ultimately their intentions to vaccinate, ‘Access’ looks at the ability of a caregiver to access and afford the direct and indirect costs to seek vaccination services when and where they are offered, while ‘Readiness’ looks at the health system capacity – including workforce, supplies, and infrastructure – to provide vaccination services for communities who seek them.

In **Define,** we conducted rapid analysis using Miro© virtual whiteboard to develop archetypes of ZD families and mapped the drivers of ZDC for each archetype. Follow-up interviews, root cause analysis, and prioritization workshops with users generated “Point of View” and “How Might We” statements. In **Dream,** we conducted rapid ideation exercises (e.g., “Crazy Eights”) that generated, grouped, and prioritized intervention ideas mapped to ZD archetypes and root causes, then translated into ‘lean’ concepts. These lean concepts simply described the solution in brief, highlighted the solutions key features, linked it to an inspiration for its conceptualization and, identified prototyping resources needed. In **Design**, we refined the lean concepts through rapid prototyping and iterative testing via virtual surveys and in-person workshops into detailed interventions ready for field-based pilot testing. Users engaged in workshops during Define, Dream, and Design phases included a representation of those previously engaged in the Discover phase -ZDC caregivers, health care workers (HCWs), community health assistants (CHAs), CHPs, sub-county health management team (SCHMT), and county health management team (CHMT) members, development partners—in the two sub-counties and, immunization experts from other PATH projects. In **Deliver** and **Adapt**, we pilot tested the co-created interventions through multiple cycles of iterations embedded in two distinct rounds of implementation.

### Gender Integration

Using ZDLA gender-integration tools, we intentionally examined how gender norms, roles, and power dynamics influenced vaccination access and decision-making. This included caregiver decision-making dynamics, HCW gender biases, and any unintended gendered effects of interventions.

### Intervention Development and Pilot Testing

The Dream phase yielded three interventions (detailed concept sheets in supplementary materials):

1. Chanjo Talks Kazini (CTK)—RN: Two 90-minute, male-focused small-group sessions at male congregation sites (beach management units, markets, workplaces). Sessions participatorily addressed vaccination benefits, schedules, the mother-child health booklet (MCHB), gender norms reflections, men’s supportive roles, and myth-busting. Between sessions, men engaged CHPs, their family, and community members on vaccination as homework.
2. Quality Household Assessment and Sensitization (QHAS)—RN: Intensive, structured capacity building for CHPs, including simplified household visit standard operating procedures (SOPs) and a caregiver-friendly immunization pamphlet insert for the MCHB.
3. Chanjo Mashinani (CM)—TC: A targeted, low-cost outreach solution differentiating households >2 hours from facilities (mobile outreach) from those <2 hours (strengthened referral via community champions) and, incorporating affordable transport, lean vaccinator teams, and community champion networks.

### Pilot Testing and Program Monitoring

We deployed each of the three interventions to selected health facilities and community units over two four-month implementation rounds.

**CTK:** We monitored session attendance, completion, satisfaction, and engagement via attendance registers and self-administered surveys. Post-intervention focus group discussions (FGDs) with men and interviews with female partners assessed user feedback and knowledge application. In round 2 we added pre/post perceived spousal support measures (interviewer-administered female partner surveys) and post-intervention health system trust assessments. At endline we assessed caregiver trust in the health promise, vaccine promise, healthcare delivery, and vaccine delivery with 20 intervention and 20 control couples using a conversational trust tool ^9^.

**QHAS:** Pre/post training knowledge and skills were measured via self-administered surveys; pre/post practice was assessed through observed shadowing against the QHAS SOPs. We monitored the number of households visited, ZD and under-immunized children reached, and mentorship visits.

**CM:** We monitored caregiver travel distance, the number of outreach sessions conducted, number of ZDC identified and vaccinated.

We conducted ongoing cycles of reviews and iterations within each round and structured After-Action Reviews (AARs) and stakeholder dissemination workshops to inform refinements.

### Ethics and Governance

The work was integrated into NVIP’s 2024–2025 annual work plans. Maseno University Ethics and Review Committee approved all activities; informed consent was obtained from all participants. SCHMT sub-teams provided oversight and ensured health system integration.

## RESULTS

### Participants

We engaged 798 participants (464 female, 334 male) across two sites, including caregivers, CHPs and CHAs, healthcare workers, SCHMT and CHMT officials, and community leaders.

### Diagnosing the Problem: From Screening to Archetypes

In Discover, we employed differentiated screening strategies matched to each context. In RN, CHPs administered household screening tools to identify ZD and under-immunized children within community units. In TC, where populations are nomadic and settlements scattered, we combined facility-based line-listing with outreach camp screening to locate missed children.

Analysis of Discover insights generated six distinct caregiver archetypes in each site, with some shared characteristics but context-specific manifestations (See figures 3 and 4). For each archetype, we categorized associated drivers according to an archetype’s attempt to vaccination; “tried but failed” or “never tried”. In RN, emergent archetypes were: (1) disabled caregivers—physically challenged, less knowledgeable, financially disadvantaged; (2) young/teen caregivers—limited social support, less knowledgeable, facing stigma; (3) elderly caregivers—less knowledgeable, less likely to keep schedules; (4) caregivers engaged in income generation—economically capable but preoccupied (e.g., fish mongers, casual laborers); (5) caregivers who delivered at home—missed opportunities; and (6) typical caregivers—mothers with no obvious risk factors but having a ZDC.

**Figure 3.**
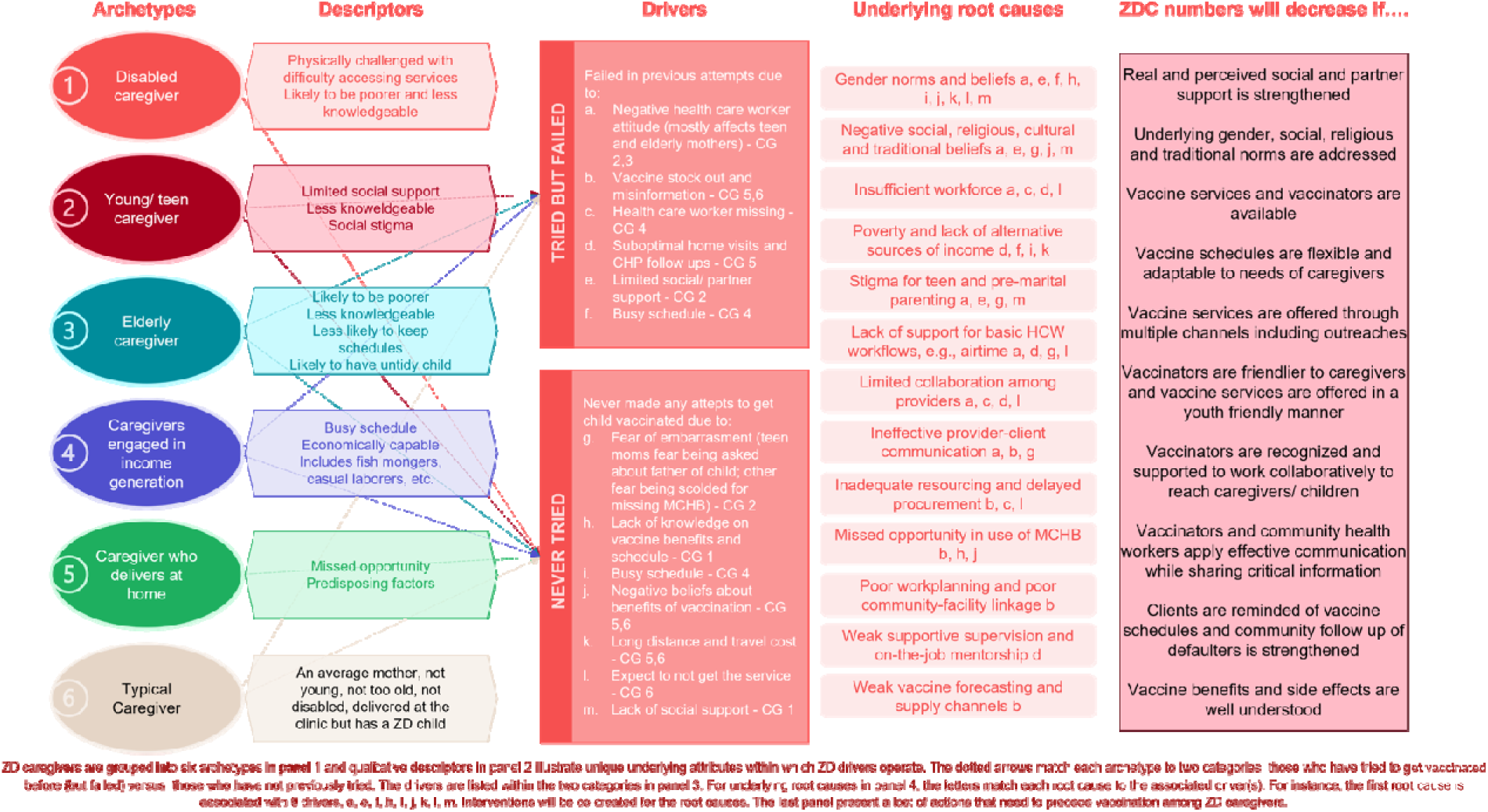
ZD archetypes in Rachuonyo North sub-county, Homabay County.

In TC, archetypes were: (1) alcoholic caregivers—involved in brewing/consumption, neglectful; (2) caregivers in remote places—less knowledgeable, facing access challenges, reliant on outreach, likely nomadic; (3) caregivers involved in farming—poor, time-constrained; (4) caregivers involved in fishing—economically capable but seasonally mobile; (5) caregivers who delivered at home—missed opportunities; and (6) typical caregivers— living near facilities, with facility-based records, but lacking knowledge to motivate immunization-seeking.

**Figure 4.**
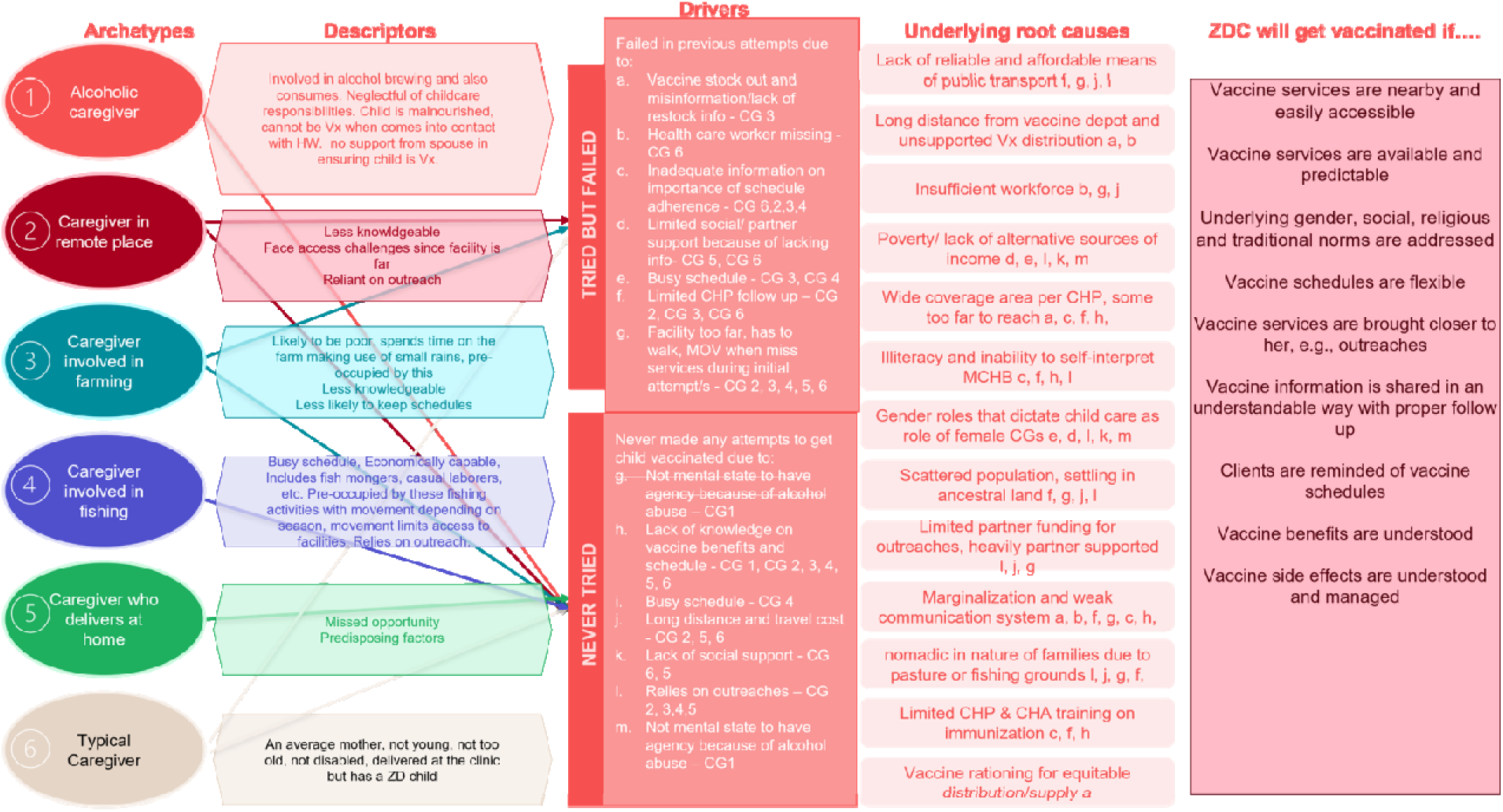
ZD archetypes in Turkana Central sub-county, Turkana County.

### Understanding Why: Priority Drivers and Root Causes

Our analysis generated 18 and 25 drivers explaining ZDC in RN and TC, respectively. Participatory prioritization workshops with stakeholders condensed these to 11 and 10 priority drivers, respectively. Following expert validation and triangulation with secondary data, we rated each driver’s evidence-strength (robust, moderate, limited, or insufficient), contribution to ZDC numbers (substantial, moderate, limited, or insufficient) and the EGH framework domains of Intent, Access and Readiness ^7^. Weighted rankings informed a prioritized list of 4 drivers per site and 15 and 10 root causes in RN and TC respectively (Table 2).

**Table 2.**
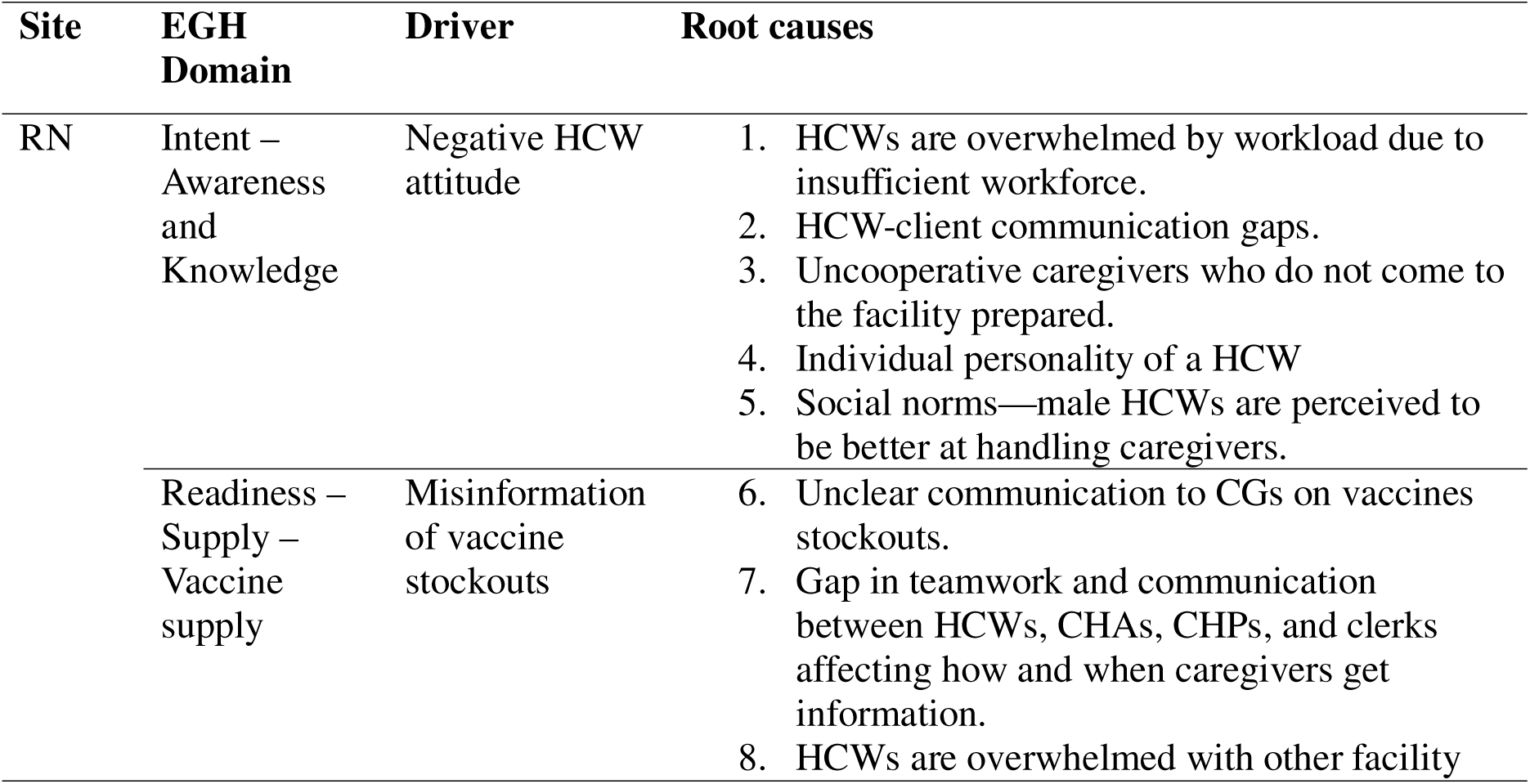
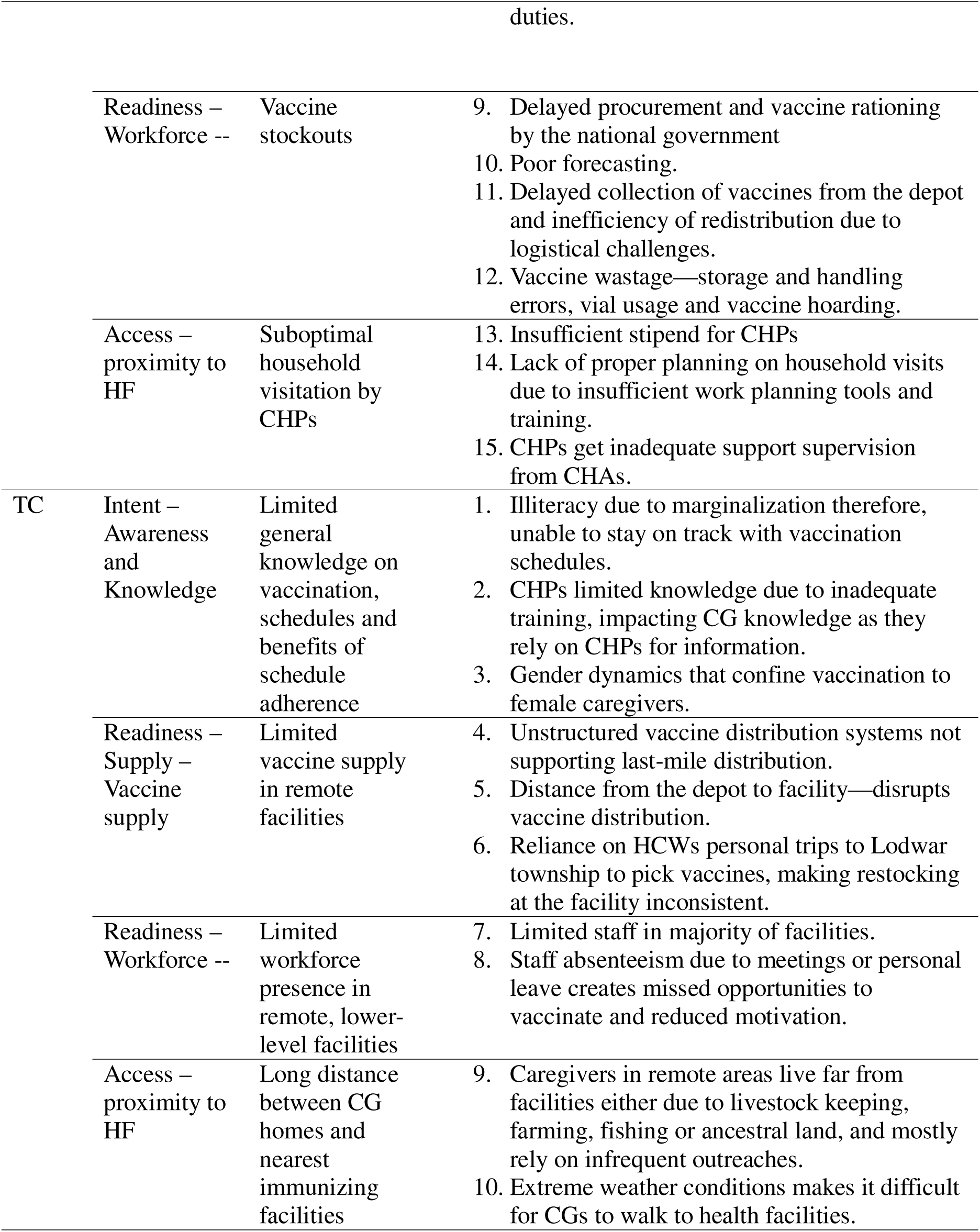
Final list of prioritized drivers and corresponding root causes across the two ZDLA sites in Kenya.

In RN, four of six archetypes had ZDC in both “tried but failed” and “never tried” profiles; two archetypes (disabled caregivers and home-delivery caregivers) had ZDC with no prior vaccination attempt. Of 13 drivers, 6 linked to “tried but failed” and 7 to “never tried” ZDC. The most emergent root causes were gender norms and beliefs (9 drivers) and negative social, religious, cultural, and traditional beliefs (5 drivers).

In TC, three of six archetypes spanned both “tried but failed” and “never tried” profiles; another three (alcoholic caregivers, fishing caregivers, and home-delivery caregivers) had ZDCs with no prior attempt. Of 14 drivers, 7 linked to each category. The most common root causes were marginalization and weak communication systems (6 drivers) and gender roles (5 drivers).

### Co-Designing Solutions: From Archetypes to Intervention Concepts

The co-creation process was deliberately inclusive, engaging ZDC caregivers, CHPs, CHAs, healthcare workers, SCHMT officials, and partner representatives. A User Advisory Group composed of users engaged during the Discover phase, provided continuous feedback across virtual surveys, in-person validation workshops, and panel discussions to test feasibility, desirability, and potential impact of emerging concepts. This iterative refinement yielded three prioritized interventions; each tightly linked to specific archetypes and root causes:

### Chanjo Talks Kazini (CTK) —Rachuonyo North

The prominence of gender norms and limited male partner support as root causes (9 drivers), combined with economic demands that kept male caregivers at workplaces rather than clinics, led to the “How Might We” question: How might we sensitize male partners to improve their immunization knowledge and strengthen their support? CTK was designed as two 90-minute, male-focused small-group sessions delivered at male congregation sites— beach management units, markets, and workplaces—rather than health facilities. Sessions participatorily addressed vaccination benefits, schedules, MCHB, gender norms reflection, men’s supportive roles, and myth-busting. Between sessions, men engaged CHPs, family, and community members on vaccination as homework.

### Quality Household Assessment and Sensitization (QHAS)—Rachuonyo North

Poor quality household visits and missed opportunities among home-delivery caregivers, disabled caregivers, and teen mothers pointed to a system-level gap: CHPs lacked structured tools to identify ZDC and tailor sensitization. The design question became: How might we improve the quality of household level assessment and sensitization? How might we create avenues for continuous professional development and tools to support household visitation workplans and improve household assessments and sensitization? QHAS was designed to deliver intensive, structured capacity building for CHPs, including simplified household visit standard operating procedures and a caregiver-friendly immunization pamphlet insert for the MCHB.

### Chanjo Mashinani (CM)—Turkana Central

In TC, access was a dominant driver. Caregivers in remote places faced walks of up to 30 km (approximately 3 hours one way) or motorcycle fares of KES 1,500 (∼USD 12) to reach the nearest facility. The “How Might We” questions focused on bringing services closer and addressing delays for home deliveries. CM was designed as a targeted, low-cost outreach solution differentiating households >2 hours from facilities (mobile outreach) from those <2 hours (strengthened referral via community champions), incorporating affordable transport, lean vaccinator teams, and community champion networks.

**Figure 4.**
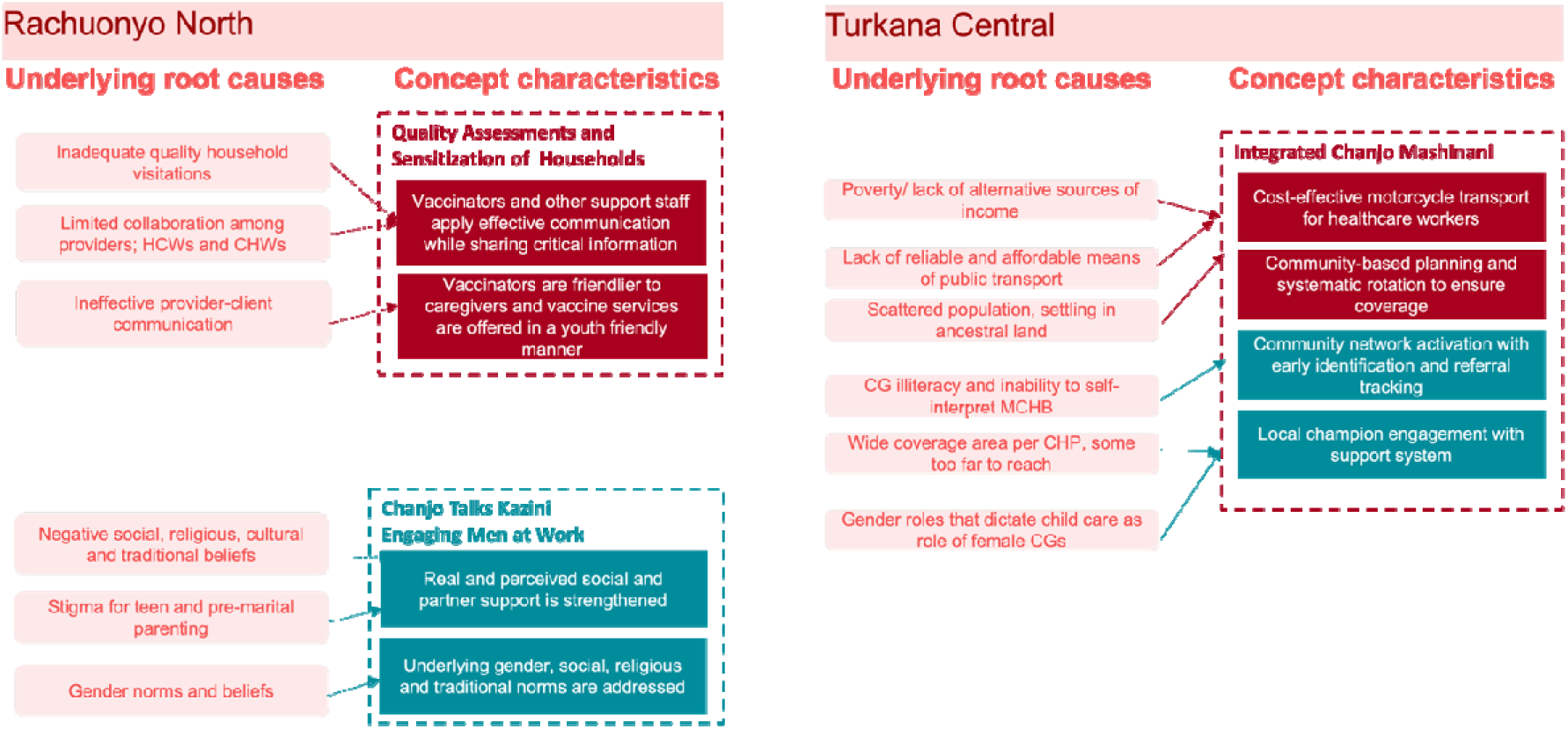
Final map linking root causes to the interventions pilot tested across the two ZDLA sites in Kenya.

### Delivering and Adapting: Implementation Cycles and Continuous Learning

We pilot-tested interventions across facilities and community units over eight months, covering two rounds of implementation, each round included multiple cycles of iterations, with SCHMTs co-creating training, monitoring, and reporting tools, identifying facilitators, and training implementers. A continuous learning (CL) model guided each intervention, led by SCHMT sub-teams and supported by structured AARs between cycles.

### Chanjo Talks Kazini

In Round 1, we established that men could be motivated to commit to vaccination-focused sessions, that learning about the MCHB was particularly inspiring, and that local health system facilitators could deliver sessions with additional training. However, post-intervention focus group discussions uncovered an unanticipated barrier: male caregivers feared HIV screening would be conducted at sessions, deterring attendance—a critical insight in Homa Bay County, which bears the highest national HIV prevalence. We also learned that female caregivers needed practical assistance and emotional support rather than symbolic male clinic attendance. In Round 2, we refined CTK to: (1) explicitly address HIV-testing fears through myth-busting and clearer mobilization messaging; (2) reorient content toward female caregivers’ actual support needs; and (3) strengthen direct linkage with QHAS to ensure household follow-through. We expanded targeting to all households with a child under one, a pregnant woman, and a male caregiver.

### Quality Household Assessment and Sensitization

Round 1 established the feasibility of delivering low-cost, targeted, practical CHP training on standard procedures for identifying ZD and UI children and sensitizing households, deployed across three villages in three priority community units. Blinded pre/post shadowing informed refinements. In Round 2, we strengthened male engagement components, improved targeting of priority households, and deepened direct linkage with CTK during household mapping. We expanded to an entire priority community unit with stronger partnership with the link health facility. As one Ministry of Health official noted, *“CHPs’ role is highly task oriented. Initiatives such as QHAS that break down CHP roles into simple step-by-step operational guidelines alongside intensive training and mentorship on those guidelines show promise in improving quality of work done by CHPs.”*

### Chanjo Mashinani

Round 1 revealed that a blanket outreach budget was insufficient for scattered settlements extending up to 70 km from facilities. We conducted 18 outreaches across six sites, but logistics required ad hoc adjustments. In Round 2, we planned outreach costs in advance with facility teams for priority villages, added two new facilities, and conducted 12 outreaches across six sites—including two uniquely remote sites requiring crossing River Kerio by foot and Lake Turkana by boat. The last time most of the sites involved had an outreach was more than a year before, while for some, it was their first time. This emphasized the need for intentional resource prioritization for extremely hard-to-reach locations, where most ZDC caregivers delivered at home resulting in weak health system linkage. We brainstormed with the CHMT and SCHMT on ways to sustain these low-cost outreaches. These included Kenya’s nationally funded health insurance scheme—Social Health Insurance Fund (SHIF)— which sustains health facilities based on the capacity of their target population, and leveraging other health partner programs and activities to deliver vaccination services in priority locations.

Referral strengthening evolved as community champions (administrative leaders, religious leaders, traditional birth attendants, and mother-to-mother support group members) linked ZDC caregivers to CHPs for referral, either by checking the MCHB to identify missed antigens or making phone calls. A facility in-charge reflected: *“We labeled our stakeholders as champions; our chief can now speak about immunization during his barazas; our admin can speak in any forum about immunization and its importance. I really love the idea of everyone coming on board to advertise this idea of children coming for immunization. It has become easier for me because everyone in the community is supporting me through this idea.”*

### Gender Dynamics: Evolving Understanding

Our understanding of gender barriers evolved significantly through implementation. In RN, initial hypotheses assumed male clinic attendance indicated support; CTK piloting revealed that practical assistance and communication mattered more. In TC, male caregivers’ months-long absence for livestock herding left female caregivers managing both household chores and income generation. This gendered labor burden interacted with distance: women could not spare time for facility visits. Our evolving understanding highlighted that distance to female caregivers’ worksites mattered as much as distance to facilities.

### Intervention Outcomes

The following outcomes represent the culmination of the diagnostic, co-design, and adaptive implementation journey described above.

### Chanjo Talks Kazini

Of 188 male caregivers enrolled, 113 (60.1%) completed both sessions. Satisfaction was high (92.5%). Ninety-five percent reported positive intentions to support vaccination, and 96% indicated increased communication about vaccination with partners, CHPs, healthcare workers, and community members. About three quarters (74.4%) of female caregivers perceived their male partners as supportive post-intervention compared to 37.1% pre-intervention. At baseline, perceived support was highest for vaccination-related decision-making (66.8%) and lowest for talking to healthcare workers (16.5%). Male caregivers’ health system trust, particularly enthusiasm for the “vaccine promise,” increased significantly versus control sites.

Qualitative interviews with male CTK graduands and their female partners revealed positive perceptions across all households. Post-intervention, men reported reading and storing the MCHB, asking questions about child health, assisting with caretaking, and accompanying wives to clinics. One participant described the intervention as providing the “fuel” (knowledge) to sustain existing care and support. Another noted: *“This intervention has reengineered our minds as men. For the first time we have been engaged directly and challenged on our traditional [roles].”* A third reflected: *“We finished the training on date 19th. The following week she was going to the clinic. I tried to understand her better this time and offered her transport to go to the clinic. I never used to do this.”*

### Quality Household Assessment and Sensitization

We trained and supported 68 CHPs and 5 CHAs. Across the two rounds, 874 priority households with children under one year were reached, identifying 15 ZD and 39 under-immunized children—all subsequently vaccinated. Blinded pre/post shadowing showed significant improvements in key sensitization and assessment process steps. Challenges included limited smartphone access for digital reporting, complex caregiver contexts requiring multiple CHP skills, vaccine stockouts, and caregiver mobility.

### Chanjo Mashinani

Across two rounds in four facilities, 30 low-cost outreach events were conducted at 12 unique sites. In total, 266 ZD children were reached. Twenty-one community champions linked ZD caregivers to CHPs for referral. Referral strengthening for “near” households (<2 hours from facility) complemented direct outreach for “far” households. Cost comparison demonstrated substantial savings over conventional outreach. Conventional outreach, requiring a 4WD vehicle from Lodwar (over 130 km to a remote health facility), up to 8 staff, and sub-county coordination cost approximately KES 792,000 per round. In contrast, the Chanjo Mashinani model using local motorcycle transport, lean teams of 4 key personnel, facility-level coordination, and targeted prioritization cost KES 139,208—yielding over 70% cost reduction while reaching priority hard-to-reach villages.

### Cross-Site Comparison

The two sites revealed shared and divergent characteristics. Gender dynamics were prominent in both but manifested differently: RN centered on male opposition and lack of support, while TC centered on gendered labor burden and male absence. Access dominated in TC but not RN, where services were geographically available but socially constrained. Intervention adoption was greater in TC, where community attitudes towards vaccination were positive leading to good turnouts during planned outreaches. In RN, two potential collaborators (DREAMS and MRITE) closed. ZD distribution differed markedly: higher, concentrated prevalence in TC versus fewer, sparsely distributed cases in RN.

## DISCUSSSION

This ZDLA study in Kenya integrated HCD with LxD to identify ZD profiles and drivers, co-create interventions, and adapt them iteratively. Findings in RN and TC revealed distinct archetypes, drivers, and positive outcomes from three co-designed interventions. Our findings demonstrate that combining HCD with LxD enables programs to correct course in response to unanticipated barriers, rather than discovering failures only at endline.

### Zero-Dose Archetypes and Drivers in Context

We identified six distinct caregiver archetypes in each sub-county, consistent with evidence that socio-economic factors, home delivery, rural residence, and maternal health-seeking behavior predict ZD status. Children from the poorest households experience nearly three times higher ZD rates than those from the richest in Kenya ^10–13^. What distinguishes our work is the granularity of these archetypes and their context-specific drivers.

In RN, gender norms contributed to male opposition and lack of support, while economic demands of semi-urban communities—particularly fish mongers and casual laborers— competed directly with health-seeking time. In TC, gendered labor burdens and male absence for livestock herding combined with severe geographic access barriers in low-density pastoralist settings, where 60% coverage and 51% home delivery compound exclusion. This distinction—socially-constrained versus geographically-constrained—underscores that ZD children share a label but not pathways to exclusion, requiring tailored responses.

### HCD and Continuous Learning: A Braided Approach

Conventional implementation research treats design and delivery as sequential phases: design, implementation, then evaluation ^14^. Our experience aligns with previous research suggesting that for complex, context-specific challenges such as reaching ZDC, design and delivery must be interwoven ^15^. Using ZD families as entry points for rapid qualitative research, we moved beyond survey-derived determinants to lived-experiences. The archetype approach provided actionable specificity—guiding intervention tailoring—while allowing cross-site comparison ^16^. This centers caregiver voices, challenging the tendency for programs to treat ZDC as passive targets rather than active participants in household decision-making ^16–19^. Integrating user perspective with technical and system perspective— strengthened by ZDLA’s emphasis on evidence quality and gender integration—prevented the pitfall of elegant designs that fail at the last mile ^20,21^.

### Deliver and Adapt: The Continuous Learning Engine

Our approach diverges from traditional implementation in the Deliver and Adapt phases. Rather than rolling out fixed protocols, we deployed interventions through SCHMT-led sub-teams who conducted structured AARs within and between cycles ^22,23^. PATH Living Labs and SCHMT officials collaboratively analyzed routine process data, with regular updates and facilitated discussions on outcomes. This created feedback loops that allowed real-time correction ^14,24^.

Three examples illustrate this engine. First, in CTK, HIV-testing fears emerged from post-Round 1 qualitative work—the kind of insights static evaluation designs miss ^25^. SCHMT facilitators adjusted mobilization messaging and session content before Round 2, transforming a potential dropout driver into an engagement opportunity. Second, in TC, conversations with female caregivers revealed that distance to worksites mattered as much as distance to facilities, reshaping our understanding of “access” in pastoralist settings. Third, blinded pre/post shadowing of CHPs under QHAS generated actionable feedback on skipped or poorly executed steps, enabling targeted mentorship rather than generic retraining ^26,27^.

These adaptations were possible because sub-county managers could act on insights without navigating prolonged approval chains ^23^. As one observer noted, pause-and-reflect workshops between cycles brought together SCHMT, CHMT, and immunization experts where *“the most valuable insights about feasibility, potential impact, and implementation strategies”* crystallized. This underscores a broader lesson: adaptive management requires not just data collection, but structured opportunities for local actors to interpret data and authorize change^17,28^.

### Intervention Insights Through a Continuous Learning Lens Chanjo Talks Kazini: When Adaptation Uncovers Hidden Barriers

CTK achieved 92.5% satisfaction and generated measurable shifts in spousal support and health system trust. The 60.1% completion rate, while modest, must be interpreted against historically low baselines for male involvement in maternal and child health across Africa ^29–31^. The 40% dropout between sessions signals that even male-friendly designs—delivered at congregation sites, addressing gender norms directly—face structural and perceptual barriers.

Most consequentially, iterative qualitative work revealed that men avoided CTK and health facilities more broadly due to fear of HIV testing. In Homa Bay County, with the highest HIV prevalence nationally, this fear is concrete. This confirms documented patterns of masculine health service avoidance in high-HIV-prevalence settings into immunization, demonstrating that HIV-related anxieties deter men from child health engagements ^32,33^. This barrier was invisible to baseline instruments; only continuous learning during delivery surfaced it.

### Quality Household Assessment and Sensitization: Task-Oriented Design for Frontline Workers

QHAS addressed a systems gap: CHP training on immunization receives less than one hour within Kenya’s standard 60-hour curriculum, and conventional training models show limited success in translating knowledge into practice ^2,34–36^. QHAS delivered targeted instruction on immunization assessment, ZD identification, and caregiver sensitization; integrated structured practice through observed shadowing; and embedded mentorship as a core component. Continuous learning proved essential for operational feedback. Blinded pre/post shadowing showed improvements in key steps but revealed persistent challenges: limited smartphone access, complex caregiver contexts, and vaccine stockouts undermining referral confidence ^37–39^. These findings underscore that frontline worker empowerment require parallel investments in supply chains, supervision, and digital infrastructure ^39–41^. Future scale-up efforts should explore leaner adaptations that preserve QHAS’s core emphasis on practice and mentorship while reducing implementation burden.

### Chanjo Mashinani: Cost-Effective Outreach Through Targeted Prioritization

The marked difference in ZDC—266 in TC versus 15 in RN among 874 priority households—reflects different challenges consistent with studies showing geographic disparity ^42,43^. In TC, success requires investment in mobile teams, transport solutions, and community champions. In RN, success demands precision in identifying scattered, socially excluded cases and addressing gendered barriers that hide them in plain sight. Neither approach transfers without substantial adaptation. CM demonstrates that reaching the last mile need not be prohibitively expensive. The CM model using local motorcycle transport, lean teams, facility-level coordination, and targeted prioritization cost KES 139,208, yielding over 70% savings, comparable to cost savings reported by similar pilot projects in Kenya ^39,44–46^. The costing results contributes to the evidence base on efficient resource allocation for reaching the last mile, an urgent consideration as Gavi and national programs face fiscal constraints ^40^.

### Methodological Contribution: What LxD Adds to Implementation Science

Our 4D HCD approach, enhanced by continuous learning, advances ZD research in three ways. First, it demonstrates that granular, context-specific diagnostics are feasible within routine health system timelines when end-users are engaged as co-investigators rather than mere respondents. Second, it shows that iterative prototyping and testing—virtual surveys, in-person workshops, shadowing assessments, and AARs—create feedback loops that enable real-time correction, as exemplified by the HIV-testing insight in CTK and the distance-to-worksite realization in TC. Third, the explicit integration of gender analysis throughout, rather than as a post-hoc equity check, revealed how gender dynamics manifest differently across contexts: as male opposition and lack of support in RN, versus gendered labor burden and male absence in TC ^47,48^.

A broader lesson concerns the architecture of learning within health systems. ZDLA enhanced our HCD approach by layering systematic gender integration, mixed-methods evidence triangulation, and technical collaboration with MoH and immunization experts. Yet the most transformative addition was adaptive management: by working through sub-county teams, we observed greater opportunities for continuous learning than is typical in centrally managed programs. For immunization policy, this suggests that investments in local learning capacity—training MoH officials in LxD/HCD tools, creating structured pause-and-reflect mechanisms, and delegating adaptation authority—may yield returns comparable to investments in intervention content itself ^36,40^.

### Key Takeaways

- Context-Specific Interventions: ZD strategies must be tailored to local archetypes and drivers rather than one-size-fits-all approaches.
- Address Male Fears: Interventions targeting men should proactively address anxieties such as fear of HIV testing through clear communication and myth-busting.
- Gender-Sensitive Programming: Recognize diverse gender dynamics—including male support, male absence, and female labor burdens—in program design.
- HCD: Methodologies like 4D HCD facilitate co-creation of relevant interventions by deeply understanding community needs.
- Continuous Learning and Adaptive Management: Structured feedback loops, AARs, and sub-county-level decision-making authority enable real-time course correction and should be institutionalized alongside intervention delivery.
- CHP Empowerment: Strengthening CHP capacity is vital for reaching priority households, but must be accompanied by investments in supply chains, supervision, and digital tools.
- Cost-Effective Outreach: Targeted, distance-stratified outreach models can achieve substantial cost savings over conventional approaches while improving reach in hard-to-access settings if invested in, consistently.

### Limitations

This work focused on two sub-counties with distinct contexts; findings may not be generalizable. The intensive LxD approach required significant time and resources; leaner adaptations may sacrifice depth for speed. We measured process outcomes and intermediate indicators; longer-term immunization coverage impact requires sustained follow-up. Premature closure of potential partner programs in RN, limited integration and scale opportunities. Finally, good-quality quantitative data were not always readily available at the local level, requiring primary data collection that added time and resource demands.

## Data Availability

All data produced in the present study are available upon reasonable request to the authors

## Notes

Funding: This work was supported by the Bill & Melinda Gates Foundation.

Conflicts of Interest: None declared.

### Competing Interest Statement

The authors have declared no competing interest.

### Author Declarations

Ethics committee of Maseno University gave ethical approval for this work

